# Forecasting the spatial spread of an Ebola epidemic in real-time: comparing predictions of mathematical models and experts

**DOI:** 10.1101/2024.03.14.24304285

**Authors:** James D Munday, Alicia Rosello, W. John Edmunds, Sebastian Funk

## Abstract

Ebola virus disease outbreaks can often be controlled, but require rapid response efforts frequently with profound operational complexities. Mathematical models can be used to support response planning, but it is unclear if models improve the prior understanding of experts.

We performed repeated surveys of Ebola response experts during an outbreak. From each expert we elicited the probability of cases exceeding four thresholds between two and 20 cases in a set of small geographical areas in the following calendar month. We compared the predictive performance of these forecasts to those of two mathematical models with different spatial interaction components.

An ensemble combining the forecasts of all experts performed similarly to the two models. Experts showed stronger bias than models forecasting two-case threshold exceedance. Experts and models both performed better when predicting exceedance of higher thresholds. The models also tended to be better at risk-ranking areas than experts.

Our results support the use of models in outbreak contexts, offering a convenient and scalable route to a quantified situational awareness, which can provide confidence in or to call into question existing advice of experts. There could be value in combining expert opinion and modelled forecasts to support the response to future outbreaks.

## Background

Following the initial emergence in 1976 in Zaire (now the Democratic Republic of the Congo, DRC)[1], epidemics of Ebola Virus Disease (EVD) have occurred, on average, every 12 - 24 months[2]. EVD is a viral haemorrhagic fever first caused by the Ebola Zaire virus (EZV), with a case fatality rate of 25-90%[3]. A major outbreak in North-Eastern provinces of DRC between 2018-2020 resulted in over 3300 reported cases and over 2100 deaths [4](Figure 1).

**Figure 1.**
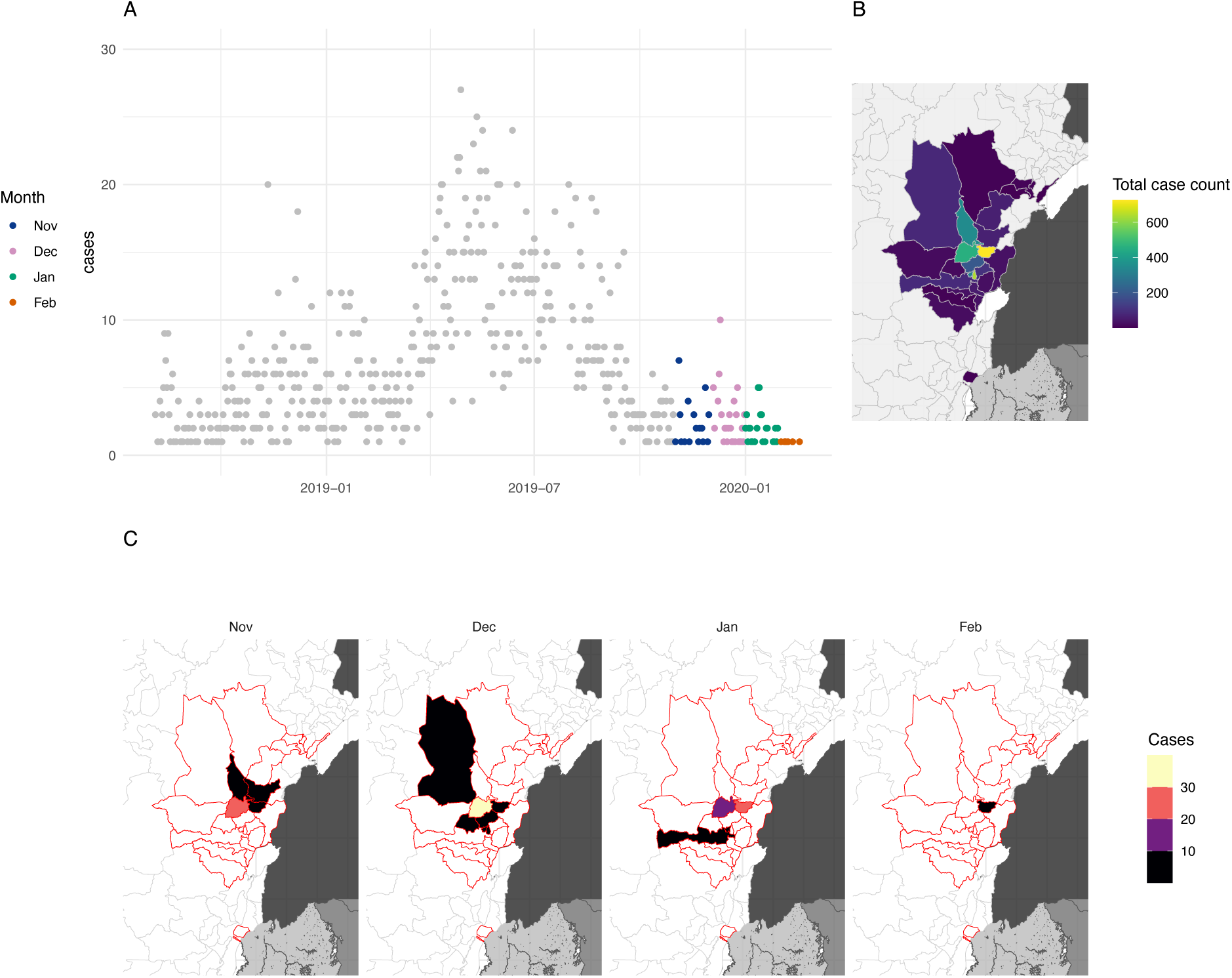
The extent of the 2018-2020 ebola outbreak in north-eastern DRC and areas included in our study. A) Daily incidence in north-eastern DRC between August 2018 and March 2020. Grey points show days prior to the study period, coloured points show days within the study period (November 2019 - March 2020), hue indicates month. B) Shows the total number of cases of ebola recorded in each Health Zone. C) Number of cases in each month and health zone during the period covered by this study, health zones outlined in red show all health zones affected by the entire epidemic.

Transmission of EZV occurs mainly through direct contact during the symptomatic phase of infection; therefore, isolation of infected individuals with strict infection control, contact tracing, and safe burials have been key to controlling past EVD outbreaks[5], although setting specific challenges can hamper containment efforts [6]. More recently, vaccination has also become a tool for outbreak control, with two vaccines now licensed for use[7][8].

EVD outbreaks typically occur in resource-poor settings where limited communication and poor accessibility make logistics of surveillance and vaccination campaigns challenging. Understanding the spatial risk of future spread is therefore useful to allow response teams to focus efforts on high-risk areas. Mathematical and statistical models have been used extensively to forecast the spread of infectious diseases, including EVD[9]. Such models rely on a combination of statistical inference based on epidemiological data and information about the mechanisms underlying the dynamics of infection. However, the dynamics of EVD are frequently governed by changing contextual factors which are challenging to forecast quantitatively. For example, violent conflicts or flooding can seriously hinder, interrupt, or even reverse the impact of containment efforts [6,10]. Moreover, changes in healthcare capacity and health seeking behaviour of patients can strengthen or weaken efforts to reduce transmission [11]. The timing and impact of these factors is notoriously difficult to predict using mathematical models.

Models are used by epidemic response experts to support decision making in the field. In addition to models, experts also make judgments as to the future spread of the virus based on their interpretation of the current status of the outbreak combined with their knowledge of other less tangible factors such as the geography, climate (eg. seasonal variation in accessibility of particular areas) and soft intelligence about the escalation of conflict in areas which may as a result, be harder to access by response teams. There are clear costs and benefits to human-made and modelled-based forecasts. Whereas models are objectively based on observations of the past outbreak dynamics and current case data, experts have additional knowledge of the complex factors surrounding the outbreak response. It is therefore difficult to assess the impact mathematical models have on decision making, how much modelled forecasts differ from those made by experts in the field and whether either modelled or human forecasts are systematically more accurate or useful. Moreover, the knowledge of experts in the field of EVD epidemiology, with a good understanding of the geographical area of study may provide an invaluable resource that is currently underused in forecasting.

Previous studies have aimed to establish the relative performance of humans and models in predicting infectious disease spread in human populations, particularly in the context of acute respiratory infections such as Influenza and SARS-CoV-2. Three studies have evaluated the predictions of humans against models, explicitly. The first of these evaluated short-term forecasts and season-wide predictions of reports of influenza-like-illness (ILI) in the United States of America (USA) [12] and two studies [13,14] compared short-term forecasts of cases of and deaths from COVID-19, firstly in Germany and Poland and secondly in the United Kingdom (UK). All three studies found that humans tended to perform better than the mathematical and statistical models selected for comparison when predicting cases. However, the COVID- focussed studies found the human ensembles performed worse than the ensemble prediction of the models when predicting deaths - These results were maintained when only self-declared ‘experts’ were included in the forecasts. A number of other studies recorded expert predictions without comparison to mathematical models. A study conducted early in the COVID-19 pandemic [15] evaluated the relative ability of laypeople and experts to predict the course of the UK epidemic over the first calendar year. The study found that both experts and laypeople typically under-predicted the impact overall, however experts’ forecasts were more accurate and better calibrated than laypeople. A study of expert predictions in the United States of America [16] evaluated their weekly forecasts of case incidence and total deaths in the first year against a pooled ensemble of all predictions. The study found that the ensemble outperformed every expert individually over the period of the study. A similar study surveyed experts regarding the total number of cases and deaths from MPox in the USA during 2022 [17], however these predictions are yet to be evaluated. Overall, these studies provide evidence that human predictions can play a valuable role in epidemiological prediction, providing a comparator and complementary method to mathematical and statistical modelling.

In this paper we extend the use of expert forecasters to predict spatial risk of transmission in the context of a local outbreak. We made monthly forecasts of the geographic spread of Ebola Virus Disease (EVD) from November 2019 to March 2020 during the declining phase of the 2018-2020 outbreak in the Democratic Republic of the Congo (DRC) using both expert predictions collected through regular interviews and with two spatially explicit computational transmission models with different spatial interaction assumptions: a gravity model and an adjacency model (where transmission can only occur between contiguous regions), see the methods section for details. Alongside supporting situational awareness, these forecasts were motivated by an aim to inform site selection for a planned vaccine trial. The objective was to identify areas that had seen no cases yet and thus were not already being supported by vaccination and other interventions, but were at high risk of still becoming affected by the EVD outbreak, thus allowing estimation of efficacy [18]. Here we evaluate the performance of the forecasts and select ensembles of the methods in predicting continued transmission and flare-ups of EVD in health zones (HZs) close to the affected area. We further study variation in forecast quality against a selection of factors related to local demography, case history and forecast implementation.

## Methods

### Expert elicitation

Experts in EVD epidemiology with knowledge of the local geography speaking English or French were identified originally by convenience sampling. The pool of experts was then expanded through recommendation from the identified experts (snowball sampling). This approach was best suited to capture the expertise of individuals who were most often temporarily based in the field.

A pilot study was carried out in November 2019. Subsequently, monthly interviews were held over WhatsApp in December 2019, January 2020, February 2020, and March 2020. All interviews were scripted. The main biases of this type of study (availability bias, representativeness bias, overconfidence, motivational bias, anchoring on past estimates) were briefly discussed during the first interview.

Experts also were provided with an interactive map of the outbreak area and surrounding health zones (HZ) showing the number of total cases during the outbreak and during the two preceding weeks for reference (supplementary figure S1). HZ were numbered to facilitate communication with the experts.

Experts were asked to estimate the number of reported probable and confirmed cases they would expect per HZ during the following month using the online MATCH Uncertainty Elicitation Tool [19] (supplementary figure S1). Through this platform, the experts and the researcher (AR) interacted in real time. The “roulette” (chips and bins) method was used.

Experts were instructed to place a total of 20 chips over the available bins (0-1 cases, 2-4 cases, …, 48-50 cases). Therefore, each chip represented for the expert a 5% probability that the number of cases was in the bin where the chip was placed. This process aimed to capture the uncertainty surrounding the expert’s estimates.

The experts were asked to estimate the number of reported cases they would expect in the HZ where there had been 1 or more cases in the 2 preceding weeks, as well as Goma (Figure 2).

**Figure 2.**
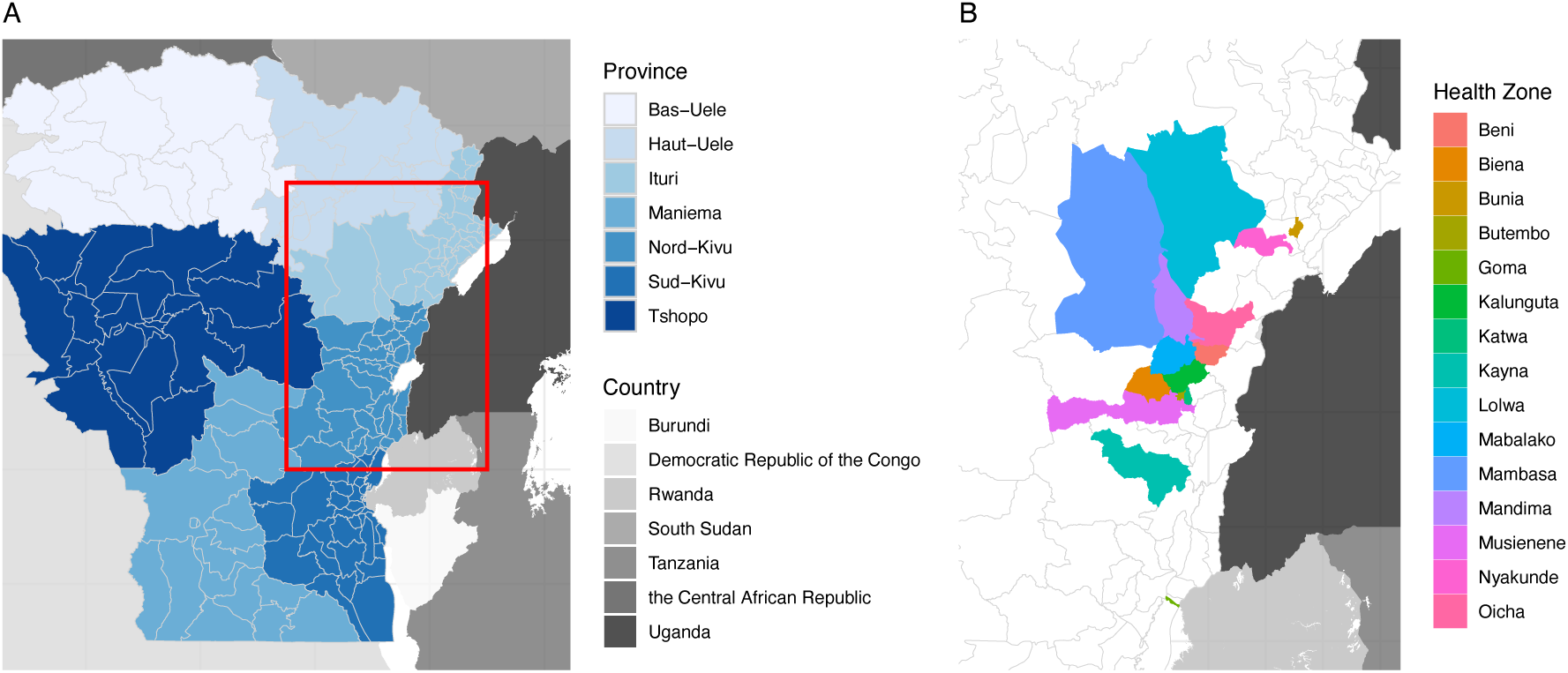
Health zones included in the model and Expert Elicitation survey. A shows the provinces around the affected area, and included in the transmission model, the red box shows the area detailed in panel B. B shows the health zones included at least once in the Expert Elicitation survey we conducted.

The experts were then asked to identify any additional HZ where they would predict 1 or more cases during the following month with >5% probability, and to also estimate the number of reported cases they would expect in these HZ. In the pilot study, carried out in October 2019, experts were asked to forecast the number of cases they expected during November 2019 in 10 HZs: Beni, Goma, Kalunguta, Katwa, Lolwa, Mabalako, Mambasa, Mandima, Nyankunde, and Oicha.

### Ethics

LSHTM ethics approval was obtained for this study (reference: 17633). Signed informed consent was taken from experts willing to participate and their verbal consent was requested again at the beginning of each elicitation.

### Modelling framework

In parallel with the expert elicitation programme, we developed a modelling framework to forecast spatial risk of infection. In the framework, incidence of cases is forecast in each Health Zone based on historical case reports. The model was formed of two components, the autoregressive component, and the spatial component.

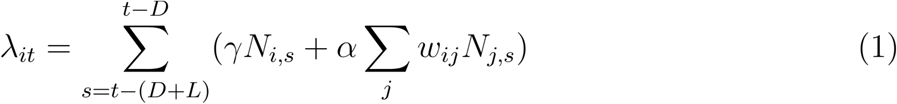

The auto-regressive component modelled the rate of infections in a particular health zone i, on day t, to be proportional to the number of cases in the same health zone (i) between dates t- (D+L) and t-D, where L is the estimated latent period and D is the estimated infectious period. The spatial component accounts for transmission between health zones, where rate of infection was proportional to the cases in each other Health Zone (i.e. ∀j j≠i) and moderated by a pairwise specific factor defined by a spatial kernel *w_ij_* We used two spatial kernels, both of which use proximity of health zones to each other and their respective population size, *P_i_* and *P_j_*. Firstly, the gravity model which treats interaction in an analogous way to Newtonian gravity with population size in place of mass, such that interaction reduces distance, *d*, raised to a power, *k*.

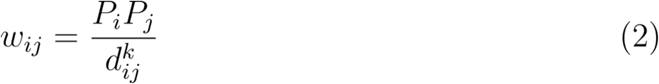

Secondly, we applied a model with adjacency-based interaction. In this model only adjacent HZs can interact. The strength of interaction between HZs is proportional to the product of their population sizes.

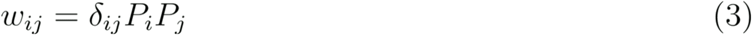

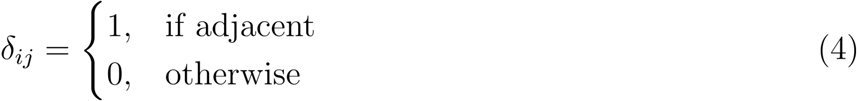

Cases were modelled as Poisson distributed such that:

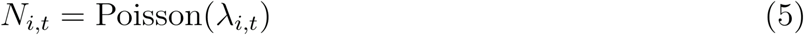

To forecast cases, we fitted the spatiotemporal model to historical data from the 60 days prior to the date the forecast was made, accounting for cases in health zones in seven regions (169 HZs) centred on the location of the epidemic; Nord-Kivu, Ituri, Tshopo, Maniema, Sud-Kivu, Haut-Uele, Bas-Uele. We fit the model using the No U-Turn Sampling (NUTS) method for Hamiltonian Monte Carlo with Stan [20], a probabilistic programming framework. We estimated *α* and *γ*, which vary the contribution of within-health-zone and between-health-zone transmission. We also estimated k, which determines how rapidly transmission rate decays with distance in the spatial component of the model. We sampled parameters from the resultant joint posterior distribution to simulate daily incidence in all HZs in the seven regions, up to and including the last day of the following month. We performed 1000 iterations for each forecast date. We then extracted the full distribution of the number of cases incident within the calendar month of interest. Forecasts were made using data up to the last day of the month prior to the forecast period.

### Enslemble forecasts

Ensemble forecasts were calculated as an average of the probabilities attributed by the members of the ensemble. For the expert ensemble the arithmetic mean was calculated across all experts with equal weighting. Similarly the model ensemble used the unweighted mean of the model forecasts. For the mixed (model and expert) ensemble, the mean was weighted such that the combined weight of the experts forecasts and the combined weight of the models forecasts were equal.

### Quantification of risk and forecast evaluation

To compare the model and the expert forecasts and score them according to the eventual true number of cases we calculated the probability attributed to cases over four thresholds, >=2, >=6, >=10 and >=20 cases.

We evaluated the forecasts using the Brier Score, a proper scoring rule which quantifies how accurate a forecast or a group of forecasts are when compared to true data after the event. The Brier score, BS, is defined as the square of the difference between the probability of observing an event and the observation *o_i_* status, which takes a value 1 or 0 for cases observed and none observed respectively. We calculated this for multiple (N) forecasts by taking the mean of the individual forecast scores.

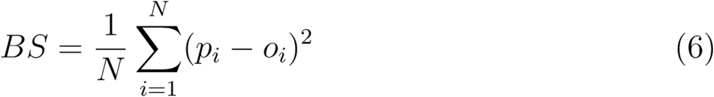

We also quantified the general bias and calibration of the forecasts by considering the hazard rate predicted by each forecast, which we calculated as the sum of probabilities attributed to exceeding each threshold. This gives the number of HZs the forecast ‘expected’ to cross the threshold in each month. To quantify the bias of each set of predictions, we took the difference between the hazard rate and the actual number of HZs that exceeded each threshold in each month. We refer to this as the *hazard gap* (HG).

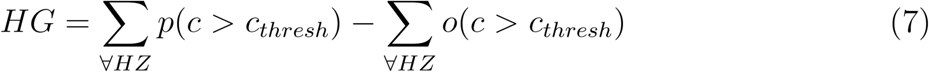

Where *p*(*c*>*cthresh*) isthe probability of threshold exceedance and *o*(*c*>*cthresh*) is the observation status of the threshold exceedence 1 if true 0 if false.

## Results

### Expert panel and health zones included in survey

Over the study period, we conducted a total of 40 interviews with 15 experts, three of which took place during the pilot phase (November 2019). Figure 3 shows the timeline of the expert elicitations.

**Figure 3.**
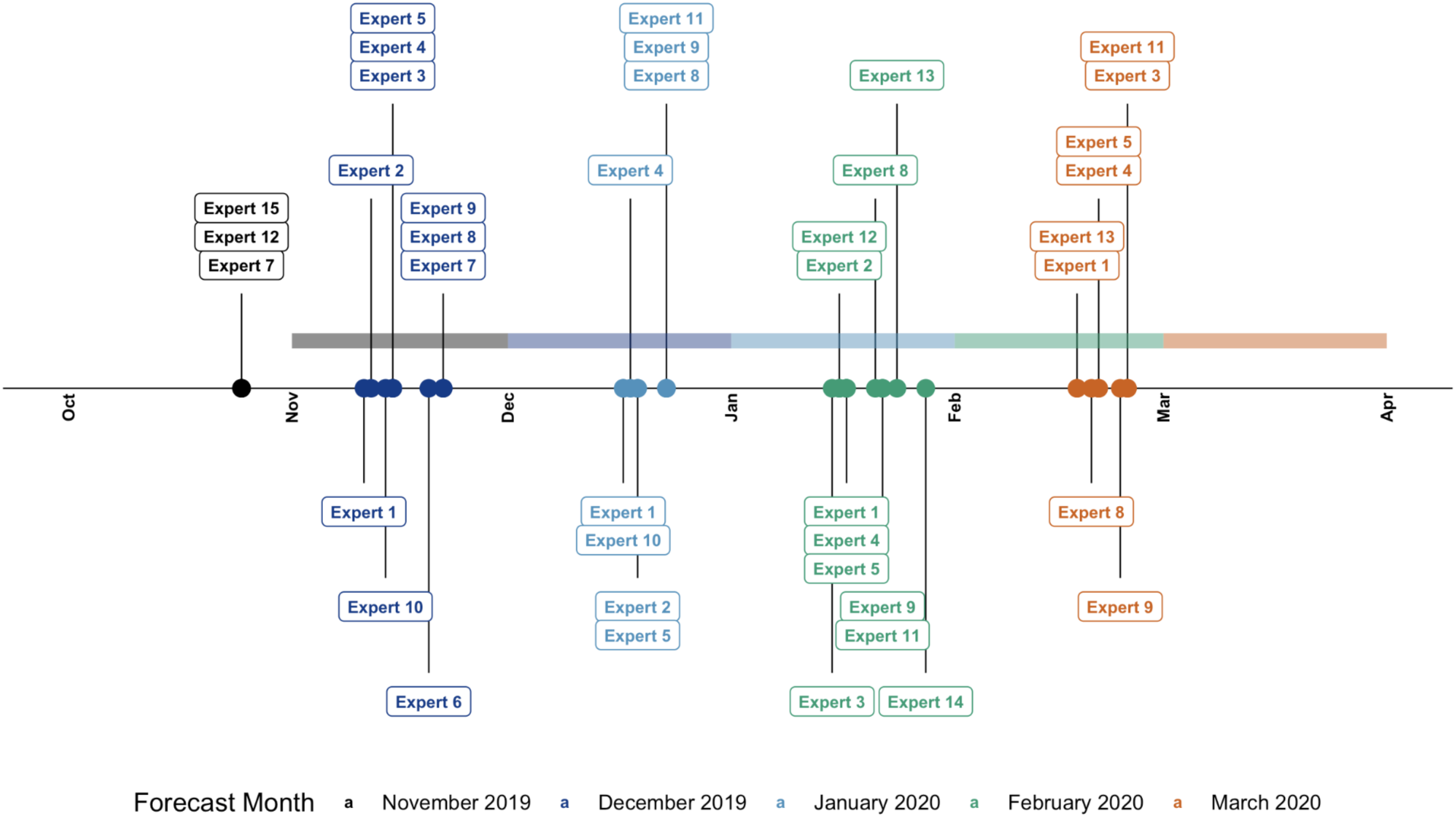
Timeline of the expert elicitation. Each point shows the date of the interview of the expert labelled to obtain forecasts for the following month. Colour indicates the month for which the forecast was made, the forecast windows are highlighted with a shaded band of the same colour.

Eight experts worked at the World Health Organization, four at the London School of Hygiene & Tropical Medicine, two for Médecins Sans Frontières, and one at the DRC Ministry of Health.

Most experts (10/15) had more than five years of experience working in infectious disease epidemiology. About half of the interviews (21 of 40) were conducted with experts that were in the outbreak area (defined HZs affected by EVD or Goma, the site of the international response base) or had been there within 2 weeks of the interview. Four experts had never been in the outbreak area.

Between December 2024 and March 2025, eight to ten experts provided monthly forecasts for 4 to 11 health zones (HZs), estimating the probability that reported cases would exceed various thresholds (2, 6, and 20 cases). Expert predictions showed variable accuracy across months and zones.

Experts correctly identified Mabalako as the highest-risk HZ in December. They attributed an average 82% probability of exceeding 2 cases; Mabalako reported 38 cases that month, exceeding all thresholds, although the probability assigned to exceeding the higher thresholds was similar to that of Beni (3 cases). Other zones with moderate cases—Kalanguta (5), and Mambasa (4)—were assigned lower probabilities and generally did not exceed higher case thresholds. Interestingly, Mandima, which had no reported cases, was assigned a relatively high probability (72%) of exceeding 2 cases, indicating some overestimation in this zone.

In January, Beni and Mabalako again accounted for most cases (22 and 11, respectively) and were recognized as high risk by the experts. However, experts underestimated the scale of the outbreak in Beni, assigning 0% probability to exceeding 20 cases there, despite this threshold being surpassed. Mandima continued to be forecasted as high risk, though no cases were reported. Predictions for other zones such as Oicha and Biena suggested moderate risk, but no cases were confirmed.

Of the 11 nominated HZs, only Beni reported confirmed cases in February (9), exceeding the 6 case threshold. Experts collectively assigned a 70% probability to this event. Similar probabilities assigned to Mabalako (60%), where no cases were reported, following cases in the prior two months. Several other zones were predicted to exceed 2 cases, yet reported none.

No cases were reported in any health zone during March. Experts broadly anticipated this, with only one expert assigning over 50% probability of exceeding 2 cases in any HZ. Beni had the highest average assigned risk at 33%.

**Figure 4.**
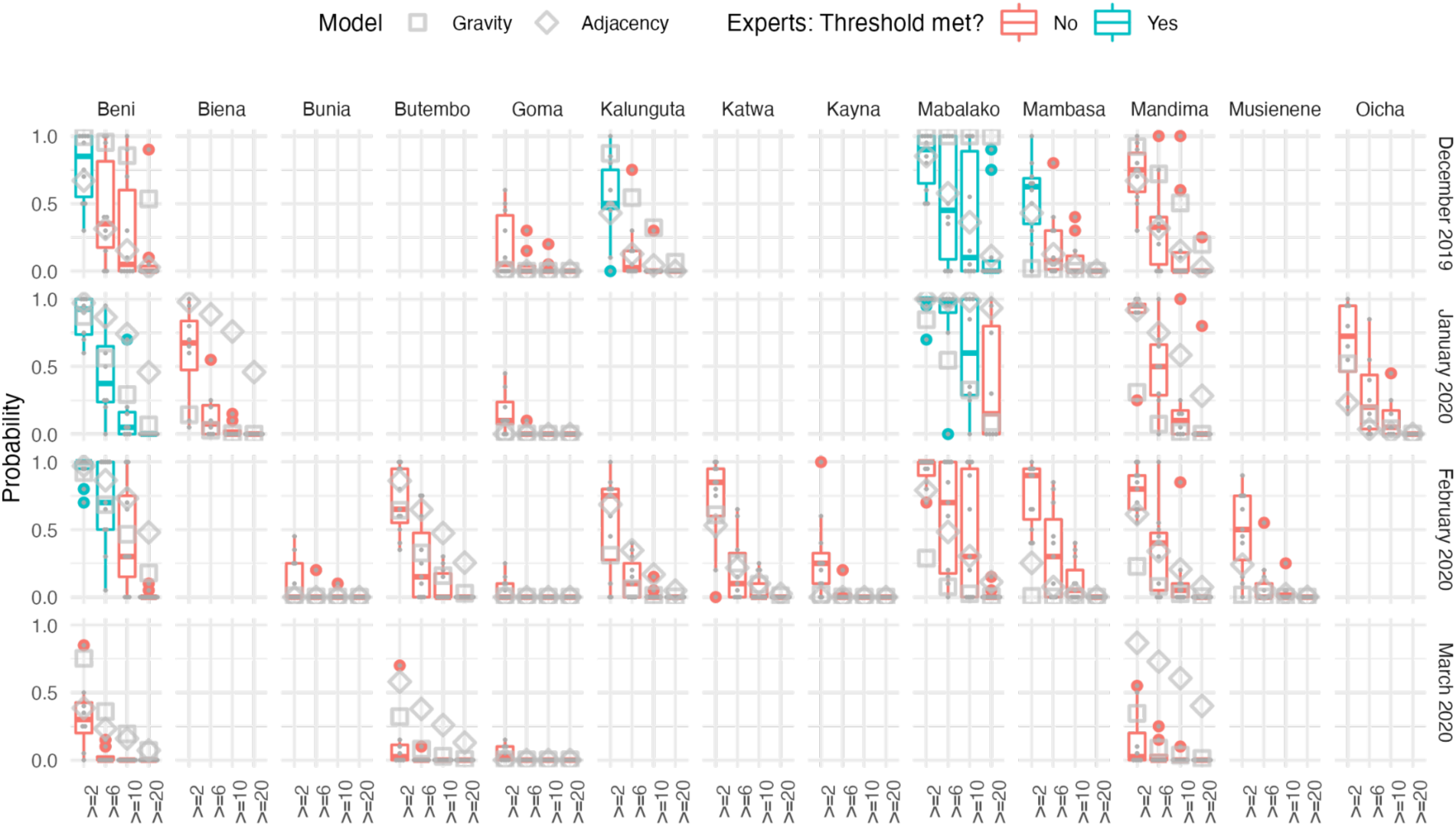
Expert elicitation results and accuracy of predictions. Only the HZs that were rated by all experts are included here. Results are shown as probabilities (vertical axes) that a given health zone (horizontal panels) exceeds a given threshold (horizontal axes) according to the experts (box plots) or models (square / diamond for gravity and adjacency models, respectively) across different months (vertical panels). Health zone / month combinations where the given thresholds were exceeded are marked in cyan, and ones where they weren’t in red.

### Performance evaluation

We evaluated the forecasts using the Brier score. The overall scores of individual experts varied between 0 and 0.6 across the four thresholds. Collectively, the experts scored best at the highest threshold (20 cases) and worst for forecasts of the lowest threshold (2 cases). The models also performed better at higher thresholds than low thresholds, but the difference was less pronounced. Overall, the gravity model ranked best amongst all forecasts at the 2 case threshold. It also ranked best for this threshold in the month of February and consistently in the top half of forecasts in December and January, however performed comparatively poorly in March, ranking higher than only one of the experts. The adjacency model also performed better than the experts overall for the 2 case threshold. Related to this, including the models improved the ensemble forecast. Although the gravity model performed better than the adjacency model for higher thresholds, together the models performed similarly to the expert ensemble forecast overall. In January and February the gravity model performed well compared to the adjacency model and the expert ensemble, however in March both models performed particularly badly compared to the experts for all thresholds. None of the experts performed consistently well relative to the others, experts 3 and 10 performed best for the 2 case threshold, whereas experts 13 and 14 did best for higher thresholds.

**Figure 5.**
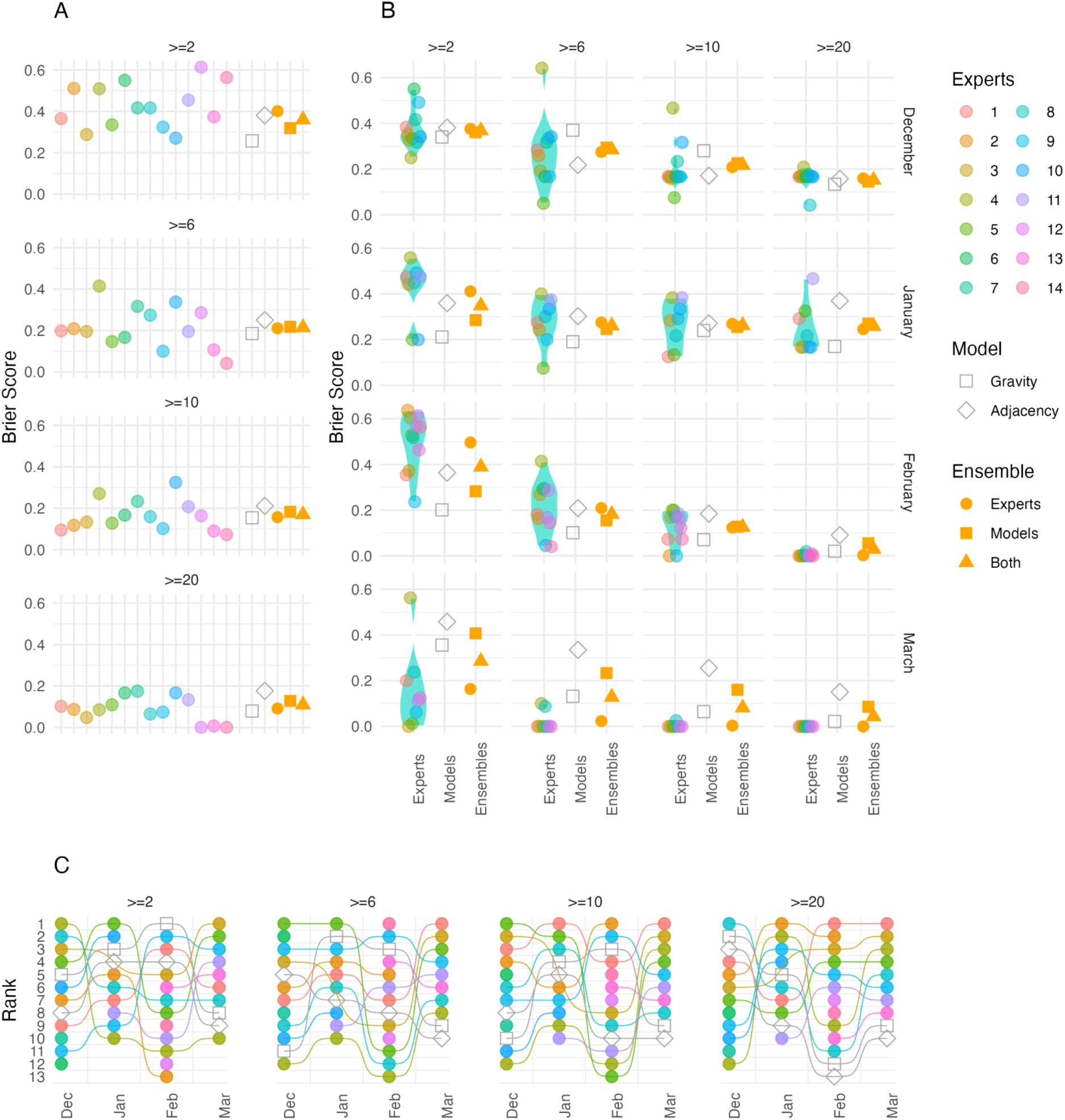
Evaluation of forecasts made by the experts, models, and ensembles. A shows the overall Brier Score for each expert, model and ensemble (calculated over all forecasts included in the study). In B each panel shows the Brier score across all health zones for each month (vertical) at each case threshold (Horizontal). Coloured points show each expert score, the violin plot shows their distribution. The grey hollow points show the model scores, the yellow points show the ensemble scores (circles show experts alone, squares show models alone and triangles show experts and models with 50% weight given to each). C Shows the ranking of each expert and model in terms of forecast performance

To evaluate how the different forecasts may impact decision making we ranked the health zones for each month, based on the probability of exceeding each threshold of cases forecast by each ensemble and by the model alone (supplementary figure S6). In general, the model and the ensembles all ranked health zones that did reach the threshold highly. In some cases the model performed better, ranking health zones that did meet the threshold higher than the experts, specifically ranking Beni Higher than Mandima in higher thresholds (>=6 and 10 cases) for the forecast of January, where Beni ultimately had cases and Mandima did not, in that month. Considering the models separately, the gravity model performed better than the adjacency model in general, with the adjacency model occasionally performing worse than the experts when ranking the HZs. This was clearest in the forecasts of November and January.

### Bias and calibration in forecasts

We evaluated the bias in each forecast type by considering the hazard gap between forecasts and actual cases. We found that experts systematically forecasted higher risk of the lowest threshold (>=2 cases) than was warranted, but tended to forecast lower risk of exceeding the highest threshold (>=20 cases) than was borne out across all HZs (Figure 6). When calculated across all months, this bias was present in 12 of the 15 experts. The models did not show clear consistent bias in either direction.

**Figure 6.**
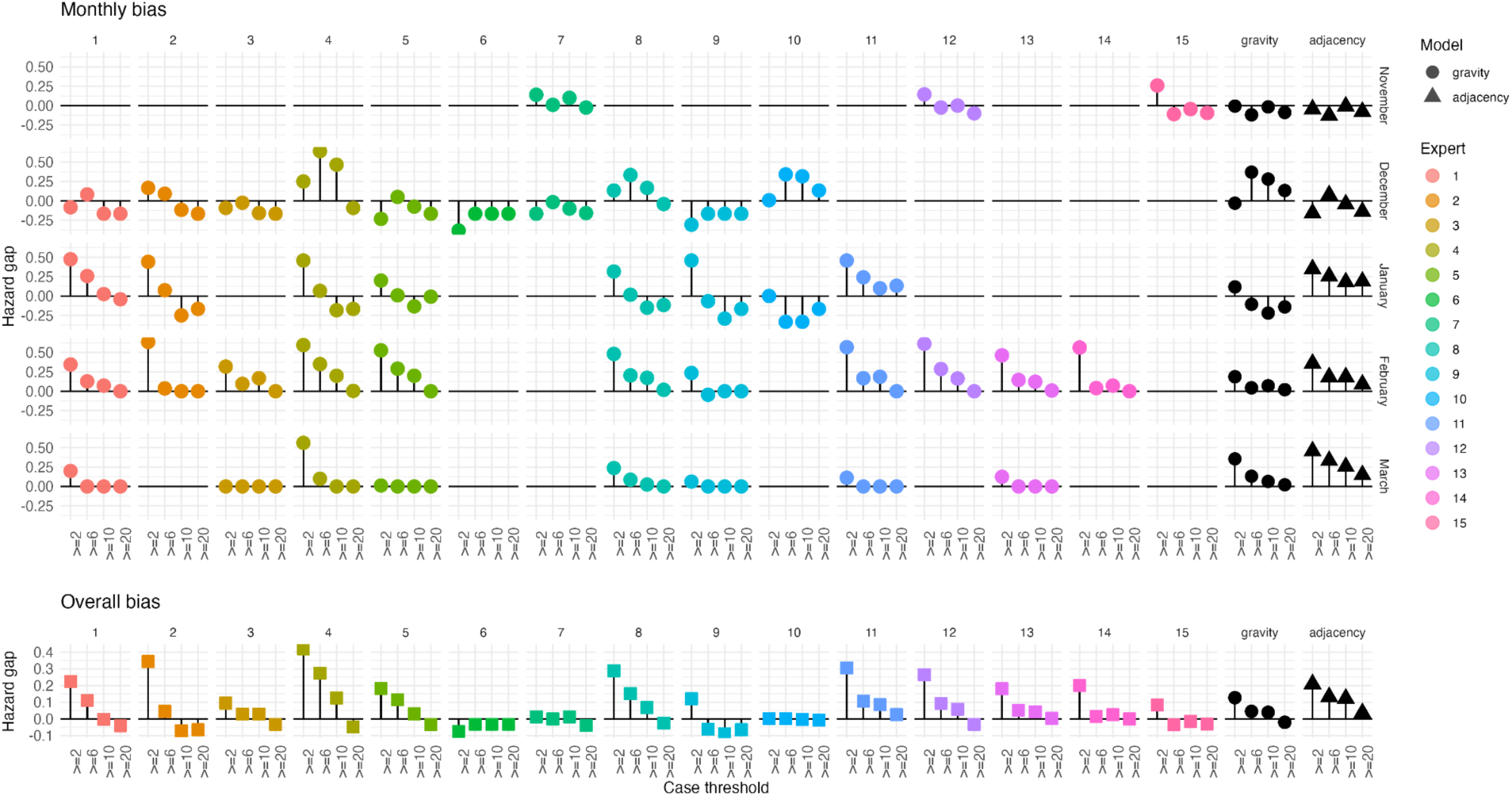
Bias and calibration of forecasts. Panels show the Hazards gap difference between the Hazard rate (expected number of exceedances across all health zones) for each threshold attributed by the forecast and the actual number of Health Zones that exceeded the associated threshold. Each panel shows one forecast (expert or model) in each month. The bottom row shows the same for each forecast calculated over the entire study period.

### Forecasting flare-ups

In addition to the health zones presented to all experts, each expert was able to nominate health zones, which they deemed at risk. Experts nominated seven further HZs to forecast in December, four in January, four in February and one in March (Table 1).

**Table 1:**
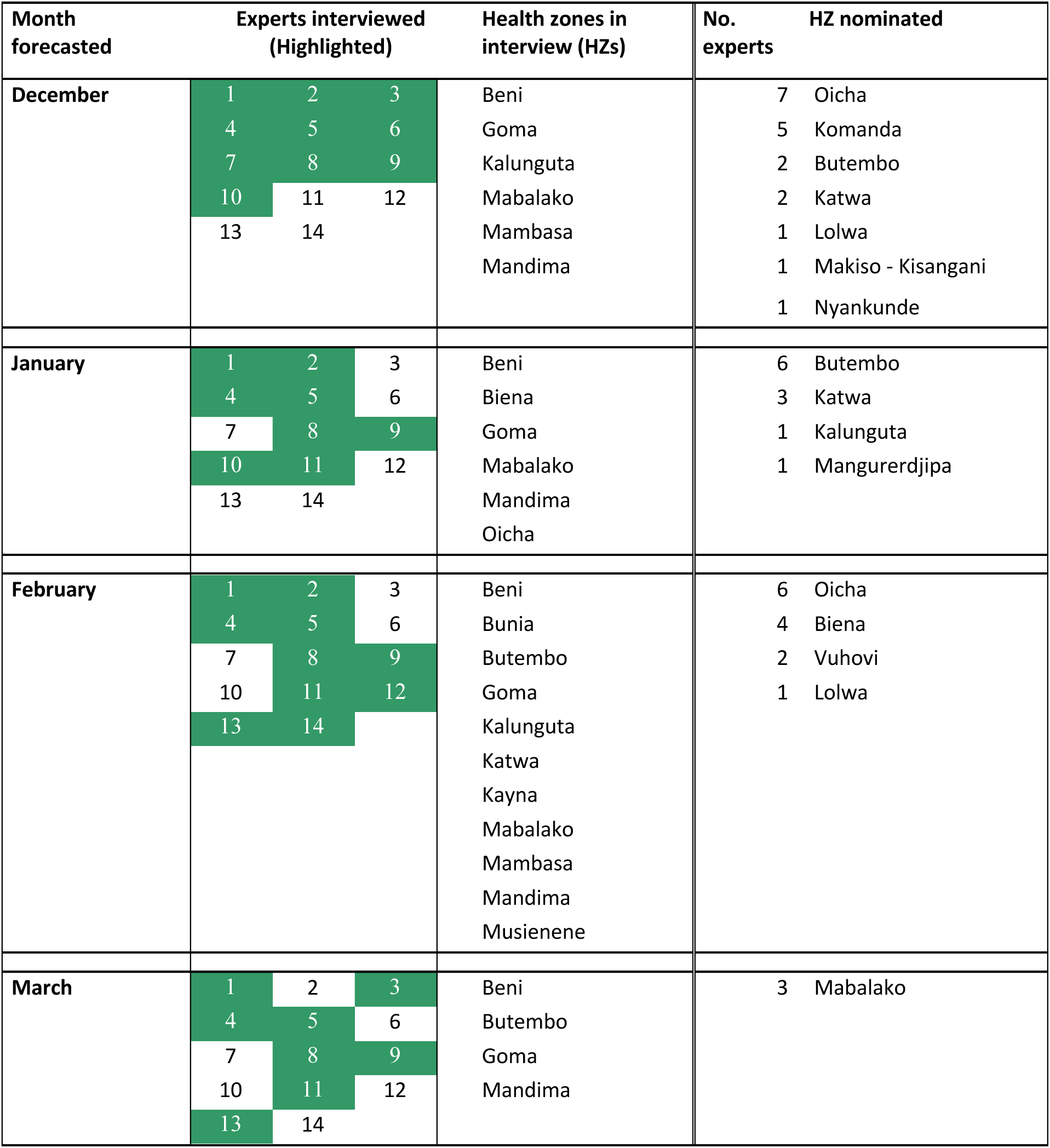
Experts and health zones included in each round of the survey. The left part of the table details the experts interviewed (highlighted in green) the health zones included in the main survey in each month. In addition, the right part of the table details the health zones nominated by experts and the number of experts that nominated each one.

The only two HZs not included in the default list but exceeding 2 cases were Butembo and Katwa. These were nominated by 2 of the 10 experts (4 and 10) both attributing 50% chance of exceeding 2 cases. In contrast, Oicha and Komanda had the most nominations with 7 and 5 each, and 6 and 4 of the 10 experts interviewed allotted greater than 5% chance of 2 or more cases in December, with attributed probabilities ranging between 5% and 95% chance of crossing the 2 case threshold.

In January, six of the eight experts interviewed nominated Butembo (1,2,4,5,9, and 11 with probabilities of 5% to 85% of crossing the 2 case threshold) while three of them also nominated Katwa (2 and 9 giving 85% and 5 giving 5%). Kalunguta and Manguredjipa were also nominated by one expert each, 5 with 10% and 11 with 50% respectively. None of the nominated health zones crossed the threshold in January.

In February six of the ten experts interviewed (2, 4, 5, 11, 12 and 14) nominated Oicha with probabilities between 10% and 95% of crossing the 2 case threshold. Four (2, 4, 5 and 11) nominated Biena with probabilities between 10% and 95% of exceedance. Experts 4 and 8 also nominated Vuhovi attributing 55% and 20% probability of threshold exceedance respectively. Expert 3 nominated Lolwa alone but gave no probability of exceeding 1 case. No HZs not included in the interview as default crossed the 2 case threshold in February.

In March three (4, 8 and 11) of the eight experts interviewed gave probabilities of 35%, 50% and 15% of exceeding the 2-case threshold respectively. No HZs not included in the interview as default crossed the 2 case threshold in March.

To compare the model with the experts we included all HZs modelled and attributed all HZs not nominated by experts an exceedance probability of 0%. To allow comparison, we also set all HZs given a probability of lower than 5% to 0% for both the gravity and adjacency models.

When considering the Brier Score (Figure 7), we found that the gravity model performed comparably to some experts when forecasting for December, and February. The adjacency model performed worse than all the experts in every month except February. In every month the ensemble of experts did better than the models and including the models in the ensembles reduced performance.

**Figure 7.**
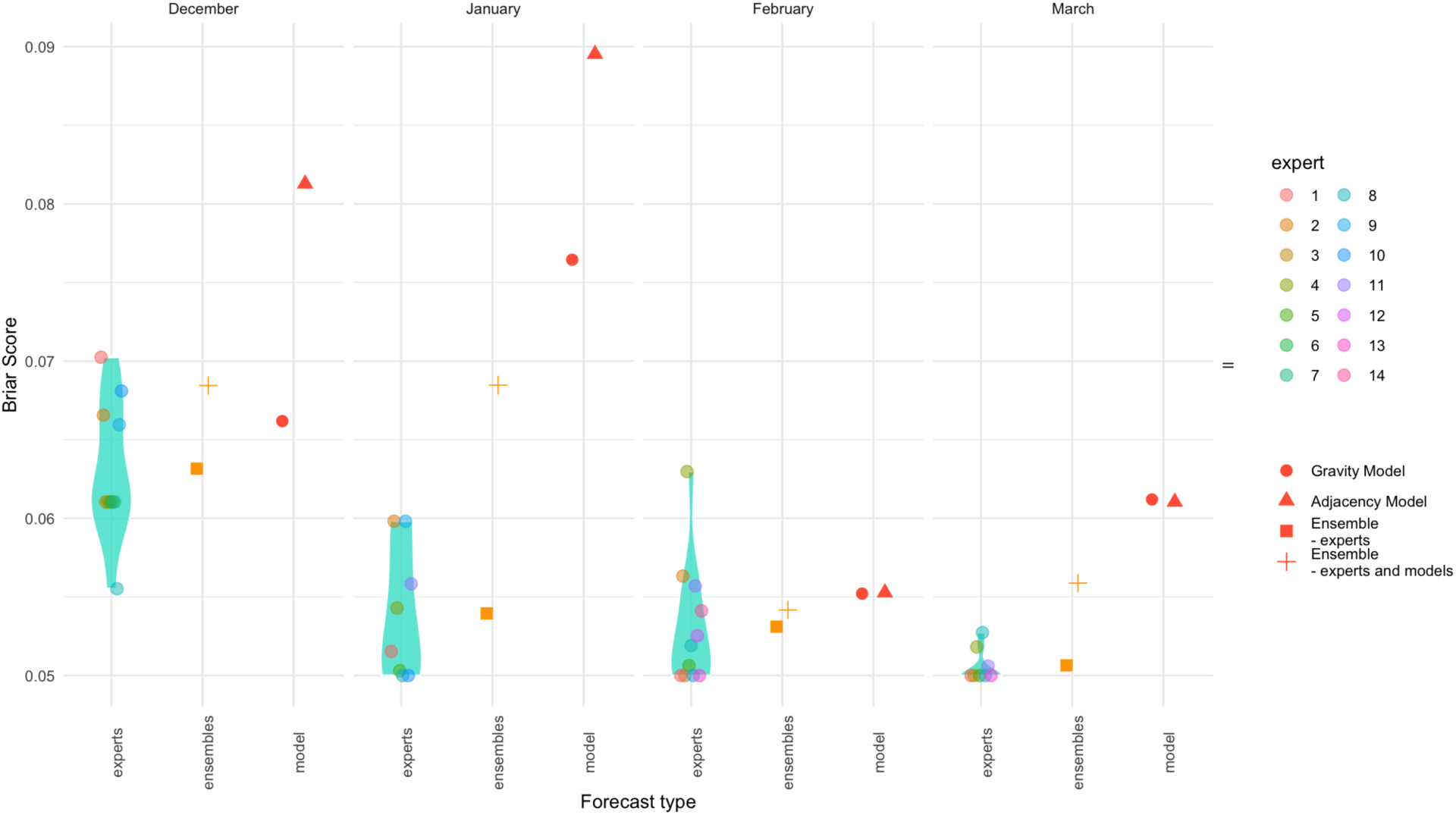
Evaluation of forecasts made in health zones not included in the main survey. Each panel (right to left) shows the Brier score across all health zones for each month. Coloured points show each expert score, density plot shows their overall distribution. The red points show the model scores, the yellow points show the ensemble scores (squares show experts alone, crosses show experts and models with 50% weight given to each).

## Discussion

We compared forecasts of the geographic spread of Ebola made by experts, with those made using a modelling framework. Since the outbreak dynamics of Ebola are highly sensitive to the changeable context in which they take place, mathematical models and expert opinions are expected to have different strengths and weaknesses, with models benefiting from objective inference from previous observations and experts able to utilise detailed knowledge about the outbreak and the changing surrounding context to make informed projections of risk. By interviewing experts and asking them to forecast risk in a structured way, we were able to compare the performance of their forecasts against those made with well-established modelling approaches in a quantifiable and robust way.

Overall, the forecasts made by the group of experts as a whole performed similarly to those of the model, with a few consistent exceptions. The model performed better than the experts when considering the lowest threshold in four of the five months covered by the survey, but performance was more comparable for the higher thresholds. The model also performed marginally better when ranking health zones by risk of ongoing transmission, indicating that use of a model may improve prioritisation of health zones when attributing resources.

We found that both methods performed better when considering higher case thresholds. This is likely to be due to a combination of bias in both forecast types towards under prediction of cases and the fact that there were few instances where the higher thresholds were reached.

Although individual experts frequently out-performed ensembles in individual instances, no individual expert outperformed the ensembles overall. This supports the practice of considering predictions from a range of experts over a smaller number of more specialist or experienced experts. The models tended to perform similarly to the ensemble representing more consistent performance across all forecasts.

Experts tended to be more biased than models, especially at low case thresholds with a tendency to over-predict cases to a greater degree than models. This bias reduced rapidly as the case threshold increased. This may be interpreted as over cautiousness from the experts regarding potential for geographic spread of the virus but confidence that transmission could be contained quickly. This trend reflects a pattern amongst previous introductions into new health zones earlier in the pandemic, where a small number of cases were reported, but the local outbreak was quickly stopped (Figure 1).

To our knowledge, our study is the first to record experts’ assessment of geographical risk at a local level during an epidemic and the first comparison of outbreak response experts’ predictions to those of models in real-time. Although direct comparison is not possible, our results lead to conclusions that are broadly similar to those from previous studies [12–14], however each of these studies found that ensemble expert forecasts performed better than the comparison models, whereas our study found no clear performance difference. This may suggest that experts are better at predicting simple time series than geographic distribution of cases. However, we cannot view these findings independently from the different survey designs or study contexts.

There are a number of important limitations to consider when interpreting our results. The context within which we conducted the study has important implications for interpretation. Due to the timing and logistics of setting up the questionnaire, the study only began in the closing phase of the epidemic, whereas the relative performance of experts and models may differ during different phases of the epidemic. For example, in the early phase where dynamics are driven more by infectious transmission than established response practices, or during the peak where changes in intervention strategy may be more influential. The stage of the epidemic also meant that there was a substantial trend towards ‘negative’ results (i.e. no threshold exceedance), which is likely to favour some forecasting methods over others.

Additionally, experts were not all interviewed on the same day and interviews occurred several days before the beginning of the month they were forecasting. In some cases experts were interviewed up to 2 weeks prior to the beginning of the month. This means that the information available differed both between experts and with the model, which was run considering data up to the first day of each month. This reduces our ability to compare models to experts directly, however, it could also be argued that this is a ‘built-in’ factor which represents the inherent challenge of eliciting predictions from experts. In addition, the interview process for experts was quite taxing and required a phone call - which can cause scheduling challenges during an epidemic response. It may be that other methods, with less arduous and more flexible data entry would improve responses.

Our analysis represents the comparison of expert forecasts to only two specific forecasting models. There are a great range of models that could have been applied in this context which may have differed in performance to those we used. We chose these models for convenience since we were applying them to the outbreak at the time of the interviews. It is also possible that some of the experts involved in the study had ingested results from our model, which were available through our online dashboard, or other models being used at the time.

Since our findings, like those of similar studies, suggest that models and experts perform comparably in this context, there is an argument that models have no value in informing expert decision making. It can be argued, however, that models remain useful in outbreak response.

Firstly, while the models performed similarly to the ensemble forecasts of the experts, there was no individual expert that performed consistently better than the models. Secondly, models are much more easily scaled and generalised making them simple to deploy in new contexts and to adapt as epidemics grow. Expert interviews are time-consuming and often inconvenient, especially in the context of outbreak response activities, which are characteristically fast paced. Models therefore offer a more convenient route to a quantified insight, which from our results, performs comparably to the way groups of experts may think. Finally, there are ways to combine both methods. For example, in the event that expert forecasts can be garnered, joint ensembles can capture information from both the expert and modelled forecasts. Further, we suggest that models can offer a role in aiding decision making by providing confidence in or calling into question expert advice that is being considered.

## Conclusions

Our analysis evaluated performance of experts and models when forecasting the spatial spread of Ebola, representing the first such study incorporating local geographic distribution and the first to focus on an epidemic in a resource poor setting. We found that forecasts made by experts and models performed comparably overall, but experts tended to be slightly more biased towards predicting that a small number of cases would persist. The results support the use of models in outbreak response and provide insight into how models and expert opinion could be combined when tackling future epidemics.

## Acknowledgements

We would like to thank Xavier de Radiguès, Neale Batra, Nabil Tabbal, Mathias Mossoko, Chris Jarvis, Thibaut Jombart, Denis Ardiet, Michel Van Herp, Silimane Ngoma, Olivier le Polain, Esther van Kleef, Noé Guinko, and Amy Gimma as well as 2 other experts who preferred to remain anonymous, for their participation as experts in this study. We would also like to thank David Smith, Thibaut Jombart, Chris Jarvis, Flavio Finger, and Anton Camacho for their helpful advice in conducting this survey.

## Code and data availability

All data and code used to process the expert interview responses can be found here: https://github.com/epiforecasts/Ebola-Expert-Interviews. The forecasts were performed using the EpiCastR package https://github.com/epiforecasts/EpiCastR. The code used for the analysis and scoring of the forecasts can be found here: https://github.com/epiforecasts/Ebola-Expert-Ellicitation

## Author contributions

AR and WJE conceived and designed the interviews. JDM and SF conceived and designed the modelling framework and the evaluation of forecasts. AR conducted the expert interviews and prepared the interview data for comparison. JDM implemented the model and performed the formal forecast evaluations. JDM, AR, WJE and SF interpreted the results. JDM and AR wrote the manuscript. JDM, AR, WJE and SF edited the manuscript.

