## Supplementary Materials for "Forecasting the spatial spread of an Ebola epidemic in real-time: comparing predictions of mathematical models and experts"

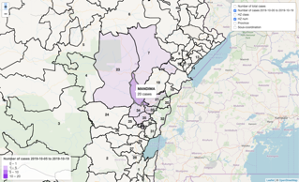

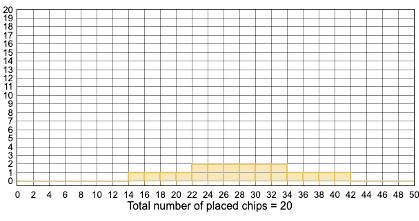

**Figure S1.** Tools used for expert elicitation. On the left, a screenshot of the interactive map provided to experts. On the right, a screenshot of the MATCH expert elicitation tool.

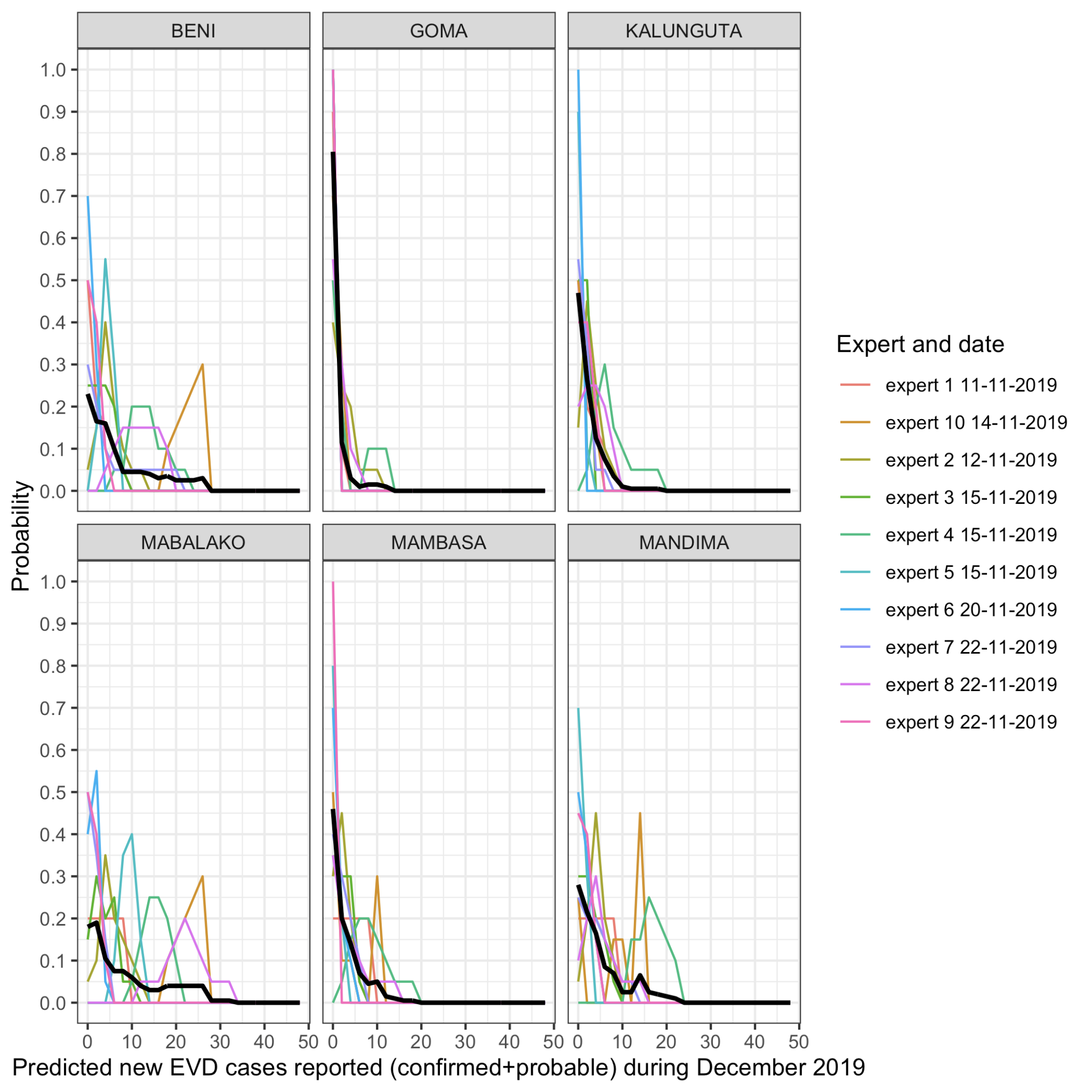

**Figure S2.** Expert forecasts made using MATCH for the distribution of cases expected in the month of December

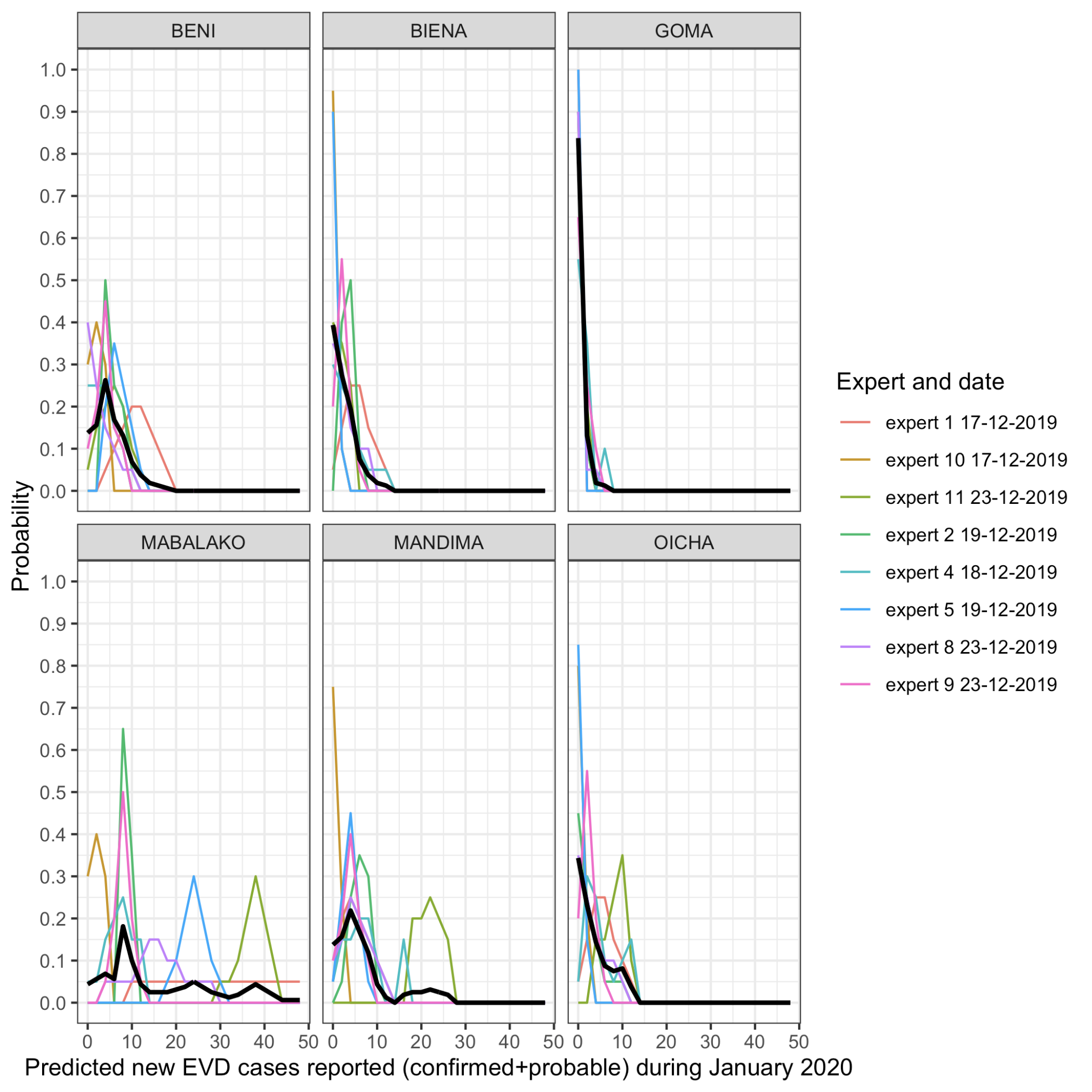

**Figure S3.** Expert forecasts made using MATCH for the distribution of cases expected in the month of January

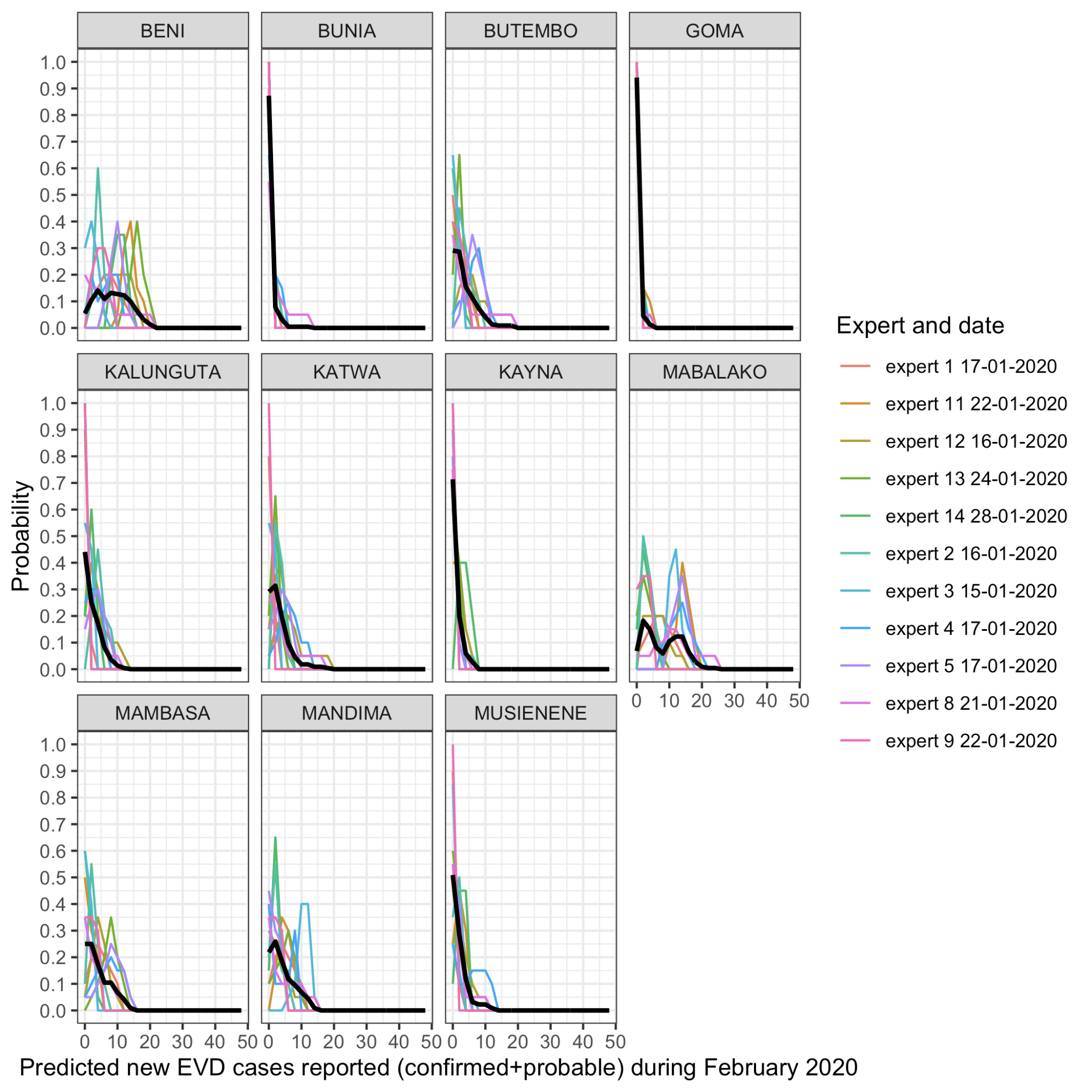

**Figure S4.** Expert forecasts made using MATCH for the distribution of cases expected in the month of February

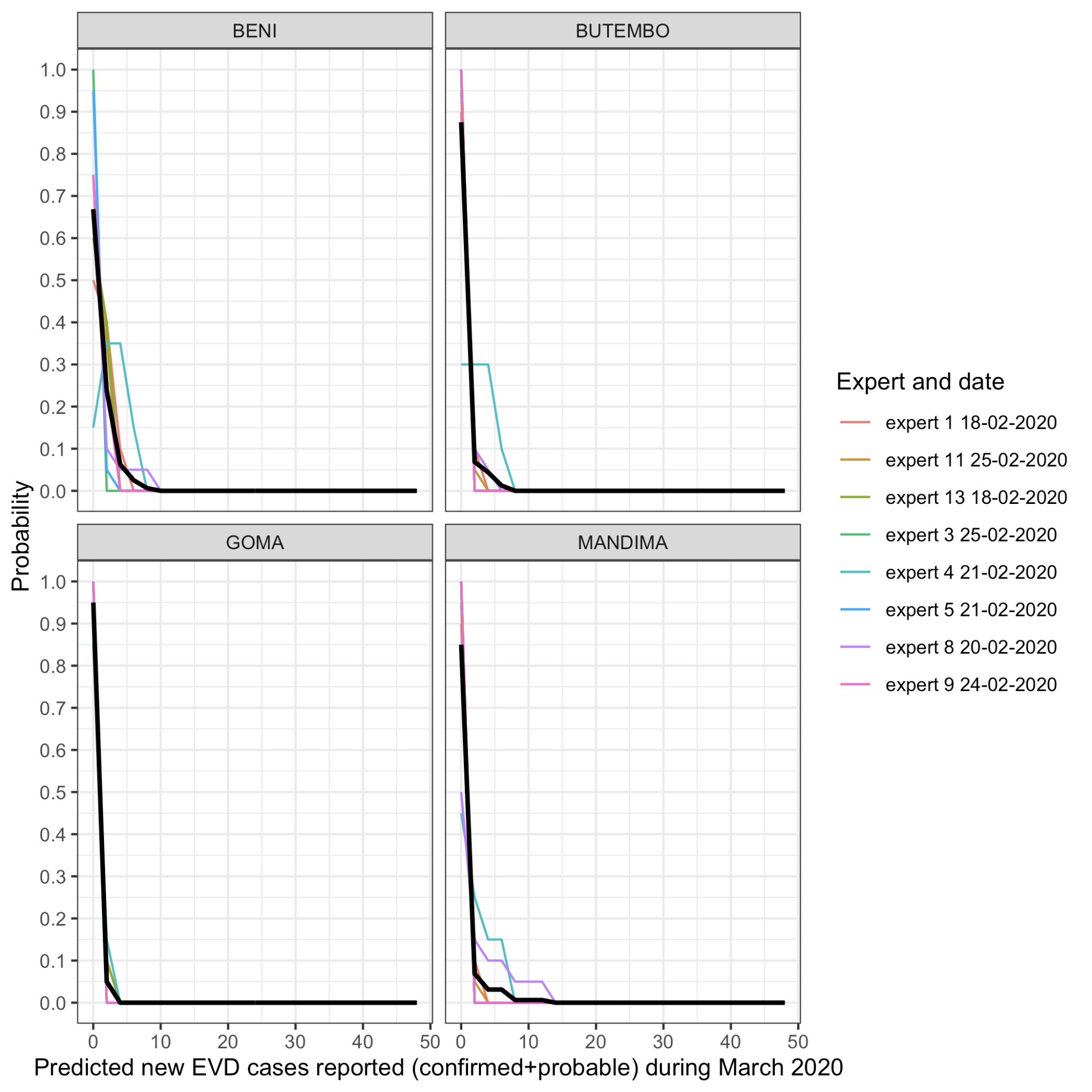

**Figure S5.** Expert forecasts made using MATCH for the distribution of cases expected in the month of March

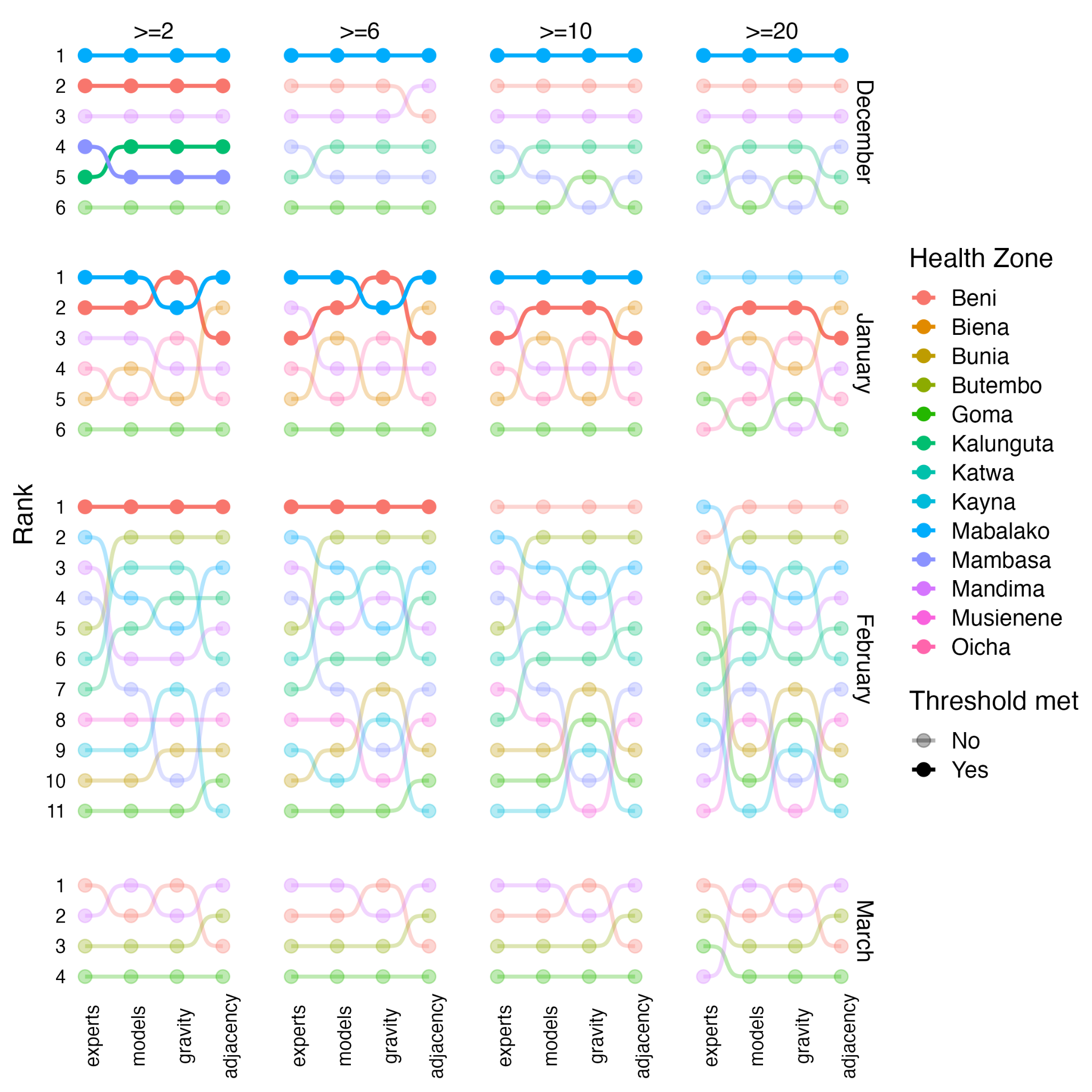

**Figure S6. Risk rank of Health Zones.** Each panel shows the rank of the Health Zones based on expert ensemble, expert and model ensemble and model forecast probabilities of exceeding each case threshold (Horizontal panels) in each month (vertical panels). Coloured lines and points indicate the Health Zone rank. Solid lines show Health Zones where the case threshold was met, faded lines show those there the case threshold was not met.
